# Dietary diversity and the risk of metabolic syndrome in a Japanese population: An analysis of baseline cross-sectional data from the J-MICC study

**DOI:** 10.1101/2024.02.15.24302906

**Authors:** Zin Wai Htay, Nobuaki Michihata, Yohko Nakamura, Yoshitaka Hippo, Jun Otonari, Hiroaki Ikezaki, Yuichiro Nishida, Chisato Shimanoe, Takashi Tamura, Mako Nagayoshi, Yasufumi Kato, Yudai Tamada, Asahi Hishida, Shiroh Tanoue, Daisaku Nishimoto, Teruhide Koyama, Etsuko Ozaki, Kiyonori Kuriki, Naoyuki Takashima, Naoko Miyagawa, Sakurako Katsuura-Kamano, Takeshi Watanabe, Kenji Wakai, Keitaro Matsuo

**Affiliations:** Cancer Prevention Center, Chiba Cancer Center Research Institute, 666-2 Nitona-cho, Chuo-ku, Chiba, 260-8717, Japan; Department of Global Health Policy, Graduate School of Medicine, The University of Tokyo, 7-3-1 Hongo, Bunkyo-ku, Tokyo, 113-0033, Japan; Department of Psychosomatic Medicine Graduate School of Medical Sciences, Kyushu University, 3-1-1 Maidashi, Higashi-ku, Fukuoka, 812-8582, Japan; Department of Comprehensive General Internal Medicine, Kyushu University Faculty of Medical Sciences, 3-1-1 Maidashi, Higashi-ku, Fukuoka, 812-8582, Japan; Department of General Internal Medicine, Kyushu University Hospital, 3-1-1 Maidashi, Higashi-ku, Fukuoka, 812-8582, Japan; Department of Preventive Medicine, Faculty of Medicine, Saga University, 5-1-1 Nabeshima, Saga, 849-8501, Japan; Department of Pharmacy, Saga University Hospital, 5-1-1 Nabeshima, Saga, 849-8501, Japan; Department of Preventive Medicine, Nagoya University Graduate School of Medicine, 65 Tsurumai-cho, Showa-ku, Nagoya, Aichi, 466-8550, Japan; Department of Public Health, Aichi Medical University, 1-1 Yasago Karimata. Nagakute, Aichi, 480-1103, Japan; Department of Epidemiology and Preventive Medicine, Kagoshima University Graduate School of Medical and Dental Sciences, 8-35-1, Sakuragaoka, Kagoshima, 890-8544, Japan; School of Health Sciences, Faculty of Medicine, Kagoshima University, 8-35-1, Sakuragaoka, Kagoshima, 890-8544, Japan; Department of Epidemiology for Community Health and Medicine, Kyoto Prefectural University of Medicine, 465 Kajii-cho, Kamigyo-ku, Kyoto, 602-8566, Japan; Laboratory of Public Health, Division of Nutritional Sciences, School of Food and Nutritional Sciences, University of Shizuoka, 52-1 Yada, Suruga-ku, Shizuoka, 422-8526, Japan; NCD Epidemiology Research Center, Shiga University of Medical Science, Seta-Tsukiwacho, Otsu, Shiga, 520-2192, Japan; Department of Preventive Medicine and Public Health, Keio University School of Medicine, 35 Shinanomachi, Shinjuku-ku, Tokyo, 160-8582, Japan; Department of Public Health, Shiga University of Medical Science, Seta Tsukiwacho, Otsu, Shiga, 520-2192, Japan; Department of Preventive Medicine, Tokushima University Graduate School of Biomedical Sciences, 3-18-15 Kuramoto-cho, Tokushima, 770-8503, Japan; Division of Cancer Epidemiology and Prevention, Aichi Cancer Center, 1-1 Kanokoden, Chikusa-ku, Nagoya, Aichi, 464-8681, Japan; Department of Cancer Epidemiology, Nagoya University Graduate School of Medicine, 65 Tsurumai-cho, Showa-ku, Nagoya, Aichi, 466-8550, Japan

**Keywords:** dietary diversity score, food variety, nutritional adequacy

## Abstract

**Background:** With the increasing burden of metabolic syndrome, it is crucial to focus on lifestyle interventions to reduce the risk. Diet is a modifiable factor that can reduce the risk of metabolic syndrome.

**Methods:** We examined the association between the dietary diversity score (DDS) and risk of metabolic syndrome using baseline data from the Japan Multi-Institutional Collaborative Cohort (J-MICC) study. In total, 75,332 participants were included in this study. A multiple logistic regression analysis was conducted to analyze the association between the DDS and metabolic syndrome.

**Results:** Inverse associations were observed between a high DDS and metabolic syndrome (adjusted odds ratio, 0.83 [95% confidential interval 0.76-0.92]). Likewise, a high DDS was associated with reduced odds of a high body mass index and hypertension. No significant associations were observed between the DDS and serum triglyceride, fasting blood glucose, or high-density lipoprotein cholesterol values.

**Conclusion:** To reduce the risk of metabolic syndrome, public health interventions should focus on promoting a diverse and balanced diet.

## Introduction

Metabolic syndrome is a cluster of interconnected non-communicable diseases, including insulin resistance, obesity, dyslipidemia, low high-density serum cholesterol, and hypertension.^1,2^ Metabolic syndrome is highly prevalent in both low- and high-income countries and is intricately linked with the incidence of cardiovascular diseases. According to the Global Burden of Disease Study 2019, stroke, cardiovascular diseases, and diabetes were the leading contributors of high disability-adjusted-life-years worldwide.^3^ Furthermore, in Japan, the age-adjusted prevalence of metabolic syndrome was reported to be 19.3% in 2005.^4^

Lifestyle modification is crucial in the management of metabolic syndrome. The dietary intake has a direct impact on metabolic risk factors. The intake of high-fiber, high-carbohydrate, and low-fat diets was reported to have positive effects on metabolic diseases.^5^ A single-center study in Japan also reported that intake of vegetable-rich diets has an inverse association with the risk of metabolic syndrome.^6^ A fast eating speed is reportedly associated with an increased risk of metabolic syndrome.^7–9^ However, previous studies have mainly explored the content and quality of the food intake, with little research into dietary diversity.

The dietary diversity score (DDS) is a composite measure of the diet quality by counting the number of food groups consumed. Consumption of diverse food groups can also result in balanced nutrient intake^10^ and thus can help reduce the risk of metabolic syndrome. The literature mainly explains the associations between DDS and the components of metabolic syndrome, such as obesity and diabetic conditions. However, inconsistent associations have been reported in different studies. Previous studies conducted among Iranian adolescents and adults reported a positive association between DDS and obesity.^11,12^ However, another study in the UK reported inverse associations between DDS and diabetic incidence^13^. Similarly, a study in Korea also stated that DDS was associated with a reduced likelihood of abdominal obesity and high triglyceride levels.^14^ Such variations in findings might be due to small sample populations and differing study designs among previously conducted studies, resulting in inconclusive evidence.

The present study therefore explored the association between the DDS and risk of metabolic syndrome in a Japanese population.

## Methods

### Study setting and participants

Our study analyzed baseline data from the Japan Multi-institutional Collaborative Cohort (J-MICC) study (version.20210901 dataset). Baseline surveys were collected during 2005-2014 at 14 different sites in Japan, including the Aichi Cancer Center, Chiba, Kanagawa, Takashima, Okazaki, Shizuoka-Daiko, Kyoto, Fukuoka, Saga, Kagoshima, Tokushima, Kyushu and Okinawa Population Study (KOPS) area, Shizuoka-Sakuragaoka, and Iga. The current study excluded the Aichi Cancer Center, Chiba, and Kanagawa data due to a lack of surveys on height, weight, blood tests, and other data necessary for defining metabolic syndrome.

Ethical approval was obtained from the ethics committee of the Aichi Cancer Center and each study site. Written informed consent was obtained from all participants. The details of the J-MICC study were reported in a previously published study profile.^15^ Participants 35-69 years old were recruited, and information was collected via a structured questionnaire. In addition, peripheral blood samples and anthropometric measurements were collected at the time of enrollment.^15^ The response rates at the study sites ranged from 19.7% to 69.8%.

### Study measurements

#### Dietary diversity assessments

Participants were administered a short food frequency questionnaire containing 47 items as part of the survey questions to obtain information on dietary habits.^16,17^ The short food frequency questionnaire assessed the food intake frequency on an 8-point scale from “not at all” to “1 to 3 or more times per day.” In this study, the food intake frequency was treated as a dichotomous variable of whether or not the participants consumed food “at least once a week”. We classified foods into five major groups (bread and grains, vegetables, meat, dairy, and fruit) based on Kant et al.^18^ To date, no universal definition of dietary diversity has been proposed. The DDS was calculated for each participant according to a counting approach that codes the intake frequency as 1 if the food group was consumed at least once a week and 0 if not.^13,19^ The DDS categorizes diets into three groups (group 3 or less, group 4, or group 5) based on the number of the five food groups (bread and grains, vegetables, meat, dairy, and fruit) consumed. To understand the intake quantity in addition to the variety, we calculated the cumulative score of five food groups based on the numbers of items consumed in each food group as the DDS-2.^20^ Each food group was assigned a maximum number of two, and the score was calculated by dividing the number of times a given item was consumed by the total number of items in each group. The obtained number was then multiplied by 2. A total score ranging from 0-10 was obtained by combining the five food groups, which were classified into four quantiles.^20^

#### Metabolic syndrome assessments

We followed the criteria of the National Cholesterol Education Program Adult Treatment Panel III (NCEP-ATP III)^21^ for the diagnosis of metabolic syndrome. Participants with biochemical data were diagnosed with metabolic syndrome if they met three of the following five conditions: 1) body mass index (BMI) ≥25 kg/m^2^; 2) systolic blood pressure ≥130 mmHg and/or diastolic blood pressure ≥85 mmHg or the use of antihypertensive medication; 3) serum triglyceride level ≥150 mg/dL; 4) serum high-density lipoprotein cholesterol (HDL-C) level <40 mg/dL in men or <50 mg/dL in women; and 5) blood fasting glucose level ≥100 mg/dL. Based on a previous study, the BMI was used instead of the waist circumference because waist circumference data were unavailable in the current study.^22^

#### Other variables

The survey questions assessed the demographic characteristics (age, sex, years of education), lifestyle factors (physical activity, history of smoking, and drinking alcohol), and family history of hypertension and diabetes mellitus. Based on previous studies on metabolic syndrome, these variables were considered possible confounders.^14,20^

### Statistical analyses

Descriptive analyses were conducted for each variable in order to understand their distribution. We also performed cross-tabulations and chi-square tests to understand the distribution of each variable with metabolic syndrome occurrence. To avoid underestimation, a multiple imputation method was applied when the number of missing observations exceeded 5% of the total observations.^23^ This method utilizes a Markov chain Monte Carlo algorithm to generate 20 complete datasets by replacing missing values with a series of predicted values. Multiple imputation assumes that the data are randomly missing. The covariates included in the imputation were the age, sex, BMI, systolic blood pressure, diastolic blood pressure, triglyceride level, HDL-C level, fasting glucose level, smoking status, alcohol intake status, family history of hypertension, family history of diabetes, DDS, and DDS-2. The input generated 20 complete datasets. The results from each input dataset were combined into a single overall estimate and standard error using Rubin’s law to reflect the uncertainty associated with missing values. This method allows for a comprehensive analysis that accounts for potential variation due to missing data.^23^

We conducted multivariable logistic regression to obtain the risk estimates of metabolic syndrome in relation to the two types of DDS. The following variables were controlled for in both models as possible confounders: age, sex, years of education, history of smoking, alcohol consumption, and physical activity. The site variable was also included in the model to account for variations in the data collection at different centers. Analyses were repeated for stratification according to metabolic syndrome risk factors.

All analyses were conducted using the Stata statistical software program (17.1; StataCorp LLC, College Station, TX, USA). The threshold level was set to 5%.

## Results

In total, 75,332 participants were included in the analysis. Table 1 shows the distribution of the participants. The majority (58.1%) of the population was ≥55 years old. More than half of the participants were women (55.3%). The average age of the study population was 55 years old. On average, 76.5% of the population engaged in physical exercise. The percentage of participants with a DDS of ≤3 was significantly higher in men (76.4%) than in women. Similar results were obtained for DDS-2, with the lowest quintile of DDS-2 predominantly comprising men (64.3%) (Supplementary Table S1).

**Table 1.** Basic characteristics of participants and distribution of the dietary diversity score.

| Variables | Total<br>N=75,332 | Dietary diversity score |  |  | p-value |
| --- | --- | --- | --- | --- | --- |
|  |  | ≤3<br>N=2,781 | 4<br>N=18,394 | 5<br>N=54,147 |  |
| Age, n (%) |  |  |  |  | <0.001 |
| 35-44 years | 12,205 (16.2) | 556 (20.0) | 4,404 (23.9) | 7,245 (13.4) |  |
| 45-54 years | 19,374 (25.7) | 820 (29.5) | 5,616 (30.5) | 12,938 (23.9) |  |
| 55-69 years | 43,753 (58.1) | 1,405 (50.5) | 8,374 (45.5) | 33,974 (62.7) |  |
| Gender, n (%) |  |  |  |  | <0.001 |
| Female | 41,694 (55.3) | 655 (23.6) | 7,832 (42.6) | 33,207 (61.3) |  |
| Male | 33,638 (44.7) | 2,126 (76.4) | 10,562 (57.4) | 20,950 (38.7) |  |
| Body mass index, mean (SD), kg/m<sup>2</sup> | 23.1 (3.3) | 23.50 (3.5) | 23.2 (3.4) | 23.0 (3.3) | <0.001 |
| Systolic blood pressure, mean (SD), mmHg | 127.8 (20.0) | 129.9 (19.8) | 127.7 (20.0) | 127.8 (20.0) | <0.001 |
| Diastolic blood pressure, mean (SD), mmHg | 78.0 (12.1) | 80.24 (12.1) | 78.5 (12.4) | 77.8 (12.0) | <0.001 |
| Triglycerides, mean (SD), mg/dL | 125.1 (98.0) | 141.10 (120.3) | 130.6 (108.3) | 122.4 (92.8) | <0.001 |
| HDL-C, mean (SD), mg/dL | 63.4 (16.6) | 59.84 (16.6) | 62.5 (16.8) | 63.9 (16.6) | <0.001 |
| Fasting glucose, mean (SD), mg/dL | 98.0 (21.0) | 100.62 (22.8) | 98.1 (21.3) | 97.8 (20.8) | <0.001 |
| Smoking status, n (%) |  |  |  |  | <0.001 |
| Current | 12,883 (17.2) | 1,024 (37.0) | 5,014 (27.4) | 6,845 (12.7) |  |
| Past | 16,455 (21.9) | 846 (30.6) | 4,749 (26.0) | 10,860 (20.1) |  |
| Never | 45,631 (60.9) | 897 (32.4) | 8,531 (46.6) | 36,203 (67.2) |  |
| Drinking status, n (%) |  |  |  |  | <0.001 |
| Current | 40,635 (54.0) | 1,942 (69.9) | 11,590 (63.1) | 27,103 (50.1) |  |
| Past | 1,925 (2.6) | 91 (3.3) | 468 (2.5) | 1,366 (2.5) |  |
| Never | 32,686 (43.4) | 746 (26.8) | 6,316 (34.4) | 25,624 (47.4) |  |
| Regular physical exercise, n (%) | 57,647 (76.5) | 1,758 (63.3) | 12,729 (69.4) | 42,692 (79.0) | <0.001 |
| Years of education, n (%) |  |  |  |  | <0.001 |
| ≤9 | 5,613 (10.4) | 373 (15.0) | 1,457 (10.7) | 3,783 (10.0) |  |
| 10-15 | 23,152 (42.9) | 1,151 (46.2) | 6,026 (44.1) | 15,975 (42.2) |  |
| ≥16 | 25,030 (46.4) | 956 (38.4) | 6,131 (44.9) | 17,943 (47.4) |  |
| Unknown | 201 (0.4) | 11 (0.4) | 52 (0.4) | 138 (0.4) |  |
| History of hypertension, n (%) | 34,146 (46.9) | 1,342 (51.2) | 8,201 (46.5) | 24,603 (46.8) | <0.001 |
| Family history, n (%) |  |  |  |  |  |
| Hypertension | 33,300 (47.7) | 1,132 (45.6) | 8,031 (47.7) | 24,137 (47.8) | <0.001 |
| Diabetes | 12,789 (18.3) | 479 (19.1) | 3,302 (19.6) | 9,008 (17.8) | <0.001 |

SD: standard deviation; HDL-C: high-density lipoprotein cholesterol

Table 2 shows the distribution of the DDS and metabolic syndrome. Overall, metabolic syndrome was reported to be the highest among subjects 55-69 years old (66.3%) and men (60.7%). The distribution of metabolic syndrome among the study sites also varied, with the highest percentage reported in Saga (17.2%) and the lowest in Tokushima (2.0%). The distribution of the DDS among the five major food groups is reported in Supplementary Table S2. In the group with a DDS ≤3, 97% and 37.6% of participants reported not eating fruits and vegetables, respectively. Similarly, participants in the lowest quantile (quantile 1) of DDS-2 reported a reduced intake of fruits (66.7%) and dairy products (15.4%).

**Table 2.** Distributions of the dietary diversity score and metabolic syndrome.

| Variables | Metabolic syndrome |  | p-value |
| --- | --- | --- | --- |
|  | Yes, n (%) | No, n (%) |  |
| Dietary diversity score |  |  | < 0.001 |
| ≤3 | 549 (4.8) | 2,232 (3.5) |  |
| 4 | 2,980 (26.2) | 15,414 (24.1) |  |
| 5 | 7,847 (69.0) | 46,310 (72.4) |  |
| Age |  |  | < 0.001 |
| 35-44 years | 1,086 (9.6) | 11,119 (17.4) |  |
| 45-54 years | 2,743 (24.1) | 16,631 (26.0) |  |
| 55-69 years | 7,547 (66.3) | 36,206 (56.6) |  |
| Gender |  |  | < 0.001 |
| Female | 4,466 (39.3) | 37,228 (58.2) |  |
| Male | 6,910 (60.7) | 26,728 (41.8) |  |
| Years of education |  |  | < 0.001 |
| ≤9 | 1,128 (14.6) | 4,485 (9.7) |  |
| 10-15 | 3,428 (44.3) | 19,724 (42.6) |  |
| ≥16 | 3,150 (40.7) | 21,880 (47.3) |  |
| Unknown | 34 (0.4) | 167 (0.4) |  |
| Smoking status |  |  | < 0.001 |
| Never | 5,734 (50.6) | 39,897 (62.7) |  |
| Past | 3,253 (28.7) | 13,202 (20.8) |  |
| Current | 2,348 (20.7) | 10,535 (16.6) |  |
| Drinking status |  |  | < 0.001 |
| Never | 4,602 (40.5) | 28,084 (44.0) |  |
| Past | 343 (3.0) | 1,582 (2.5) |  |
| Current | 6,413 (56.5) | 34,222 (53.6) |  |
| Regular physical exercise |  |  | < 0.001 |
| No | 2,931 (25.8) | 15,046 (23.6) |  |
| Yes | 8,410 (74.2) | 48,769 (76.4) |  |
| Site |  |  | < 0.001 |
| Takashima | 460 (4.0) | 3,065 (4.8) |  |
| Okazaki | 982 (8.6) | 5,568 (8.7) |  |
| Shizuoka-Daiko | 968 (8.5) | 9,189 (14.4) |  |
| Kyoto | 917 (8.1) | 5,276 (8.3) |  |
| Fukuoka | 1,444 (12.7) | 8,649 (13.5) |  |
| Saga | 1,953 (17.2) | 10,115 (15.8) |  |
| Kagoshima | 1,572 (13.8) | 6,066 (9.5) |  |
| Tokushima | 222 (2.0) | 2,218 (3.5) |  |
| KOPS area | 1,948 (17.1) | 7,782 (12.2) |  |
| Shizuoka-Sakuragaoka | 680 (6.0) | 4,837 (7.6) |  |
| Iga | 230 (2.0) | 1,191 (1.9) |  |

Table 3 presents the results of the multivariate logistic regression analysis. The total number of observations in this logistic regression model was imputed because more than 28.3% of the observations were missing for education and fasting blood glucose variables. Participants with a high DDS were significantly less likely to have metabolic syndrome than those with lower values, as evidenced by an adjusted odds ratio (aOR) of 0.83 (95% confidence interval [CI] 0.76-0.92). A similar estimate was observed for the DDS-2, with the rate of metabolic syndrome being lowest among those in the highest DDS-2 quantile, as indicated by an aOR of 0.90 (95% CI 0.82-0.99). Similarly, we found that a high DDS was associated with a reduced occurrence of a high BMI (aOR 0.91 [95% CI 0.84-0.99]), hypertension (aOR 0.84 [95% CI 0.77-0.92]), and high serum triglyceride levels (aOR 0.90 [95% CI 0.82-0.99]) (Supplementary Table S3). A similar pattern was observed for the DDS-2 and metabolic syndrome risk factors, except for high serum triglyceride levels (Supplementary Table S4). However, we found no association between the DDS and low HDL-C levels or high fasting glucose levels (Supplementary Tables S5 and S6). We also conducted stratification analyses by sex group; however, no effect modification was observed (Supplementary Tables S5 and S6).

**Table 3.**
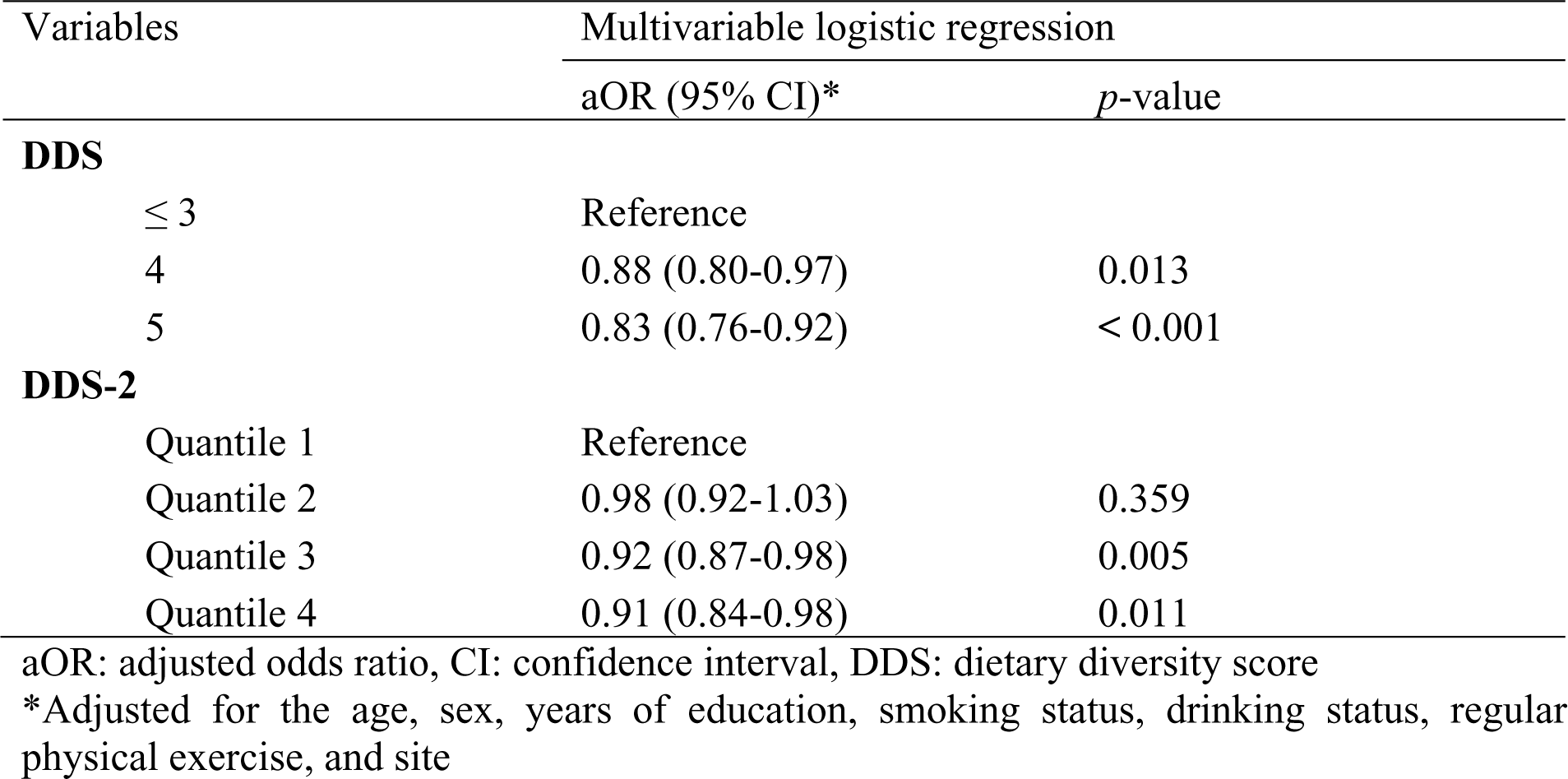
Associations between dietary diversity scores and metabolic syndrome Variables Multivariable logistic regression.

| Variables | Multivariable logistic regression |  |
| --- | --- | --- |
|  | aOR (95% CI)* | *p*-value |
| **DDS** |  |  |
| ≤ 3 | Reference |  |
| 4 | 0.88 (0.80-0.97) | 0.013 |
| 5 | 0.83 (0.76-0.92) | < 0.001 |
| **DDS-2** |  |  |
| Quantile 1 | Reference |  |
| Quantile 2 | 0.98 (0.92-1.03) | 0.359 |
| Quantile 3 | 0.92 (0.87-0.98) | 0.005 |
| Quantile 4 | 0.91 (0.84-0.98) | 0.011 |

aOR: adjusted odds ratio, CI: confidence interval, DDS: dietary diversity score
\*Adjusted for the age, sex, years of education, smoking status, drinking status, regular
physical exercise, and site

## Discussion

In this study, we collected data from various regions of Japan to examine the association between the DDS and metabolic syndrome. This is one of the few studies to focus on dietary diversity and metabolic syndrome risk factors in Japan. We found that the overall DDS had an inverse relationship with the risk of metabolic syndrome. We also found that a high DDS was associated with a reduced likelihood of reporting a high BMI, hypertension, high serum triglyceride levels, and high fasting blood glucose levels.

Both a high DDS and high DDS-2 were associated with a reduced likelihood of reporting metabolic syndrome. Our findings were consistent with those of a previous study in a Korean population, in which a lower risk of metabolic syndrome and its risk factors were reported in the higher DDS group than in the lower DDS group.^14^ This supports our hypothesis that a diverse diet is a protective factor against the risk of metabolic syndrome. The underlying assumption for the current finding is that dietary diversity is highly associated with the consumption of a balanced and adequate diet.^24^ Our study also showed that those in the diverse diet groups were more likely to consume fruits and vegetables than others. Our descriptive analyses revealed that men had a higher percentage of lower DDS scores than did women. This observation aligns with previous studies conducted in China and Australia,^25,26^ which also reported higher meat consumption among men than among women. These findings emphasize the need to prioritize the male population when promoting dietary diversity, warranting increased attention, and targeted interventions. In our study, we compared the risk estimates of the two DDS indicators, with the second indicator considering the cumulative index of dietary diversity. However, since the risk estimates were similar for both DDS indicators, we were unable to determine whether or not consuming a wide range of food groups within a single food category was particularly advantageous for health.

Analyses of metabolic syndrome risk factors also showed that a high DDS was associated with a reduced likelihood of reporting hypertension and high serum triglyceride levels. The finding that DDS is associated with a reduced likelihood of hypertension was supported by a previous study,^27^ which reported that DDS was inversely associated with serum triglyceride levels and systolic blood pressure. In addition to the carbohydrate intake, the meat intake was also reported to be associated with the risk of peripheral atherosclerosis.^28^ Furthermore, low-carbohydrate diets and calorie restriction have been reported to be effective interventions in reducing serum triglyceride levels.^29^ Similar to our findings, a previous Japanese study reported that greater dietary diversity was associated with the more frequent consumption of fiber in the diet,^30^ which can be a possible explanation to reduced serum triglyceride concentrations, thereby leading to a preventive effect.

In addition, we found that a higher DDS was associated with a reduced risk of reporting a high BMI. One underlying assumption is that a higher DDS is associated with a lower consumption of a carbohydrate-rich diet, which is the main factor leading to obesity.^31^ However, contrary to our findings, a previous systematic review reported no associations between the DDS and BMI.^32^ Similarly, another systematic review reported no conclusive evidence concerning the DDS and obesity associations.^19^ These diverse associations might be related to different cultural aspects of the involved participants. In our study, we investigated the associations by comparing the two DDS scores, and the results from both scores were consistent, indicating that consuming a diverse diet is advantageous for BMI management.

Our study had several strengths and limitations. This study offers a comprehensive evaluation of dietary diversity in the Japanese population by employing two different approaches to define the DDS. The findings from both approaches are comparable, revealing a consistent association between increased dietary diversity and a decreased risk of metabolic syndrome. In addition, our study incorporated a substantial sample size, which enhanced the reliability and statistical power of our findings. However, this study was an analysis of baseline cross-sectional data, and because of the nature of the cross-sectional study, we were unable to establish causal relationships. Although we controlled for educational status and basic demographic characteristics in our model, we were unable to control for the wealth index of the study population, which might have resulted in residual confounding. In addition, a self-reported questionnaire was used to collect data on dietary factors, the history of smoking, drinking, physical exercise, and the family history, so there was a possibility of inevitable misclassification or measurement error. This might have led to the underestimation of true associations, with the estimates biased towards the null hypothesis. Despite the potential for misclassification or measurement errors due to the use of self-reported questionnaires, our findings were relatively uninfluenced, as we identified significant associations.

Our findings revealed variations in site-specific risks for the association between DDS and metabolic syndrome. Further exploration of the prefectural-level economic situation and other possible influencing factors is recommended to explain such variations. This study did not explore the determinants of dietary diversity in the Japanese population, so further studies are recommended to determine policy interventions to implement and promote dietary diversity.

## Conclusion

In conclusion, our study found that consumption of a diverse range of foods was associated with a reduced risk of metabolic syndrome and its associated risk factors. To mitigate the risk of metabolic syndrome in the Japanese population, it is advisable to avoid restricting food intake to specific groups. Instead, promoting a varied and balanced diet may be recommended as a preventive measure against metabolic syndrome risk factors.

## Data Availability

The datasets analyzed during the current study are not publicly available.

## Sources of funding

This study was supported by Grants-in-Aid for Scientific Research for Priority Areas of Cancer (No.17015018), Innovative Areas (No.221S0001), and Platform of Supporting Cohort Study and Biospecimen Analysis (Japan Society for the Promotion of Science [JSPS] KAKENHI Grants No.16H06277 and 22H04923 [CoBiA]) from the Japanese Ministry of Education, Culture, Sports, Science, and Technology.

## Acknowledgements

We would like to thank the participants of the J-MICC study for providing the necessary information to conduct the current research.

## Disclosures

The authors declare no conflict of interests.

## Authorship and contributions

Conception and design: ZWH, NM1

Data acquisition: JO, HI, YN2, CS, TT, MN, YK, YT, AH, ST, DN, TK, EO, KK, NT, NM2, SK, TW, KW, KM, J-MICC members

Analysis and interpretation of data; ZWH, NM1

Drafting the manuscript: ZWH

Revising the manuscript: NM1, YN1, YH, JO, HI, YN2, CS, TT, MN, YK, YT, AH, ST, DN, TK, EO, KK, NT, NM2, SK, TW, KW, KM, J-MICC members

## List of Abbreviations

aOR: adjusted odds ratio
BMI: body mass index
DDS: dietary diversity score
J-MICC: Japan Multi-institutional Collaborative Cohort Study
KOPS area: Kyushu and Okinawa Population Study area
HDL-C: high-density lipoprotein cholesterol

## Supplementary Materials

Tables S1-S6

## Notes

### Competing Interest Statement

The authors have declared no competing interest.

### Author Declarations

Ethical approval was obtained from the ethics committee of the Aichi Cancer Center and each study site.

## References

1. Saklayen MG. The global epidemic of the metabolic syndrome. Curr Hypertens Rep. 2018;20. doi:10.1007/S11906-018-0812-Z

2. Kassi E, Pervanidou P, Kaltsas G, Chrousos G. Metabolic syndrome: definitions and controversies. BMC Med. 2011;9:1–13. doi:10.1186/1741-7015-9-48/TABLES/2

3. Abbafati C, Abbas KM, Abbasi-Kangevari M, et al. Global burden of 369 diseases and injuries in 204 countries and territories, 1990–2019: a systematic analysis for the Global Burden of Disease Study 2019. The Lancet. 2020;396(10258):1204–1222. doi:10.1016/S0140-6736(20)30925-9

4. Liu L, Miura K, Fujiyoshi A, et al. Impact of metabolic syndrome on the risk of cardiovascular disease mortality in the United States and in Japan. American Journal of Cardiology. 2014;113:84–89. doi:10.1016/j.amjcard.2013.08.042

5. Calton EK, James AP, Pannu PK, Soares MJ. Certain dietary patterns are beneficial for the metabolic syndrome: reviewing the evidence. Nutrition Research. 2014;34:559–568. doi:10.1016/J.NUTRES.2014.06.012

6. Bahari T, Uemura H, Katsuura-Kamano S, et al. Nutrient-derived dietary patterns and their association with metabolic syndrome in a Japanese population. J Epidemiol. 2018;28:194. doi:10.2188/JEA.JE20170010

7. Garcidueñas-Fimbres TE, Paz-Graniel I, Nishi SK, Salas-Salvadó J, Babio N. Eating speed, eating frequency, and their relationships with diet quality, adiposity, and metabolic syndrome, or its components. Nutrients. 2021;13. doi:10.3390/NU13051687

8. Nanri A, Miyaji N, Kochi T, Eguchi M, Kabe I, Mizoue T. Eating speed and risk of metabolic syndrome among Japanese workers: the Furukawa Nutrition and Health Study. Nutrition. 2020;78:110962. doi:10.1016/J.NUT.2020.110962

9. Zhu B, Haruyama Y, Muto T, Yamazaki T. Association between eating speed and metabolic syndrome in a three-year population-based cohort study. J Epidemiol. 2015;25:332. doi:10.2188/JEA.JE20140131

10. Kennedy GL, Pedro MR, Seghieri C, Nantel G, Brouwer I. Dietary diversity score is a useful indicator of micronutrient intake in non-breast-feeding Filipino children. J Nutr. 2007;137:472–477. doi:10.1093/JN/137.2.472

11. Karimbeiki R, Pourmasoumi M, Feizi A, et al. Higher dietary diversity score is associated with obesity: a case–control study. Public Health. 2018;157:127–134. doi:10.1016/j.puhe.2018.01.028

12. Golpour-Hamedani S, Rafie N, Pourmasoumi M, Saneei P, Safavi SM. The association between dietary diversity score and general and abdominal obesity in Iranian children and adolescents. BMC Endocr Disord. 2020;20:1–8. doi:10.1186/S12902-020-00662-W/TABLES/4

13. Conklin AI, Monsivais P, Khaw KT, Wareham NJ, Forouhi NG. Dietary diversity, diet cost, and incidence of type 2 diabetes in the United Kingdom: a prospective cohort study. PLoS Med. 2016;13. doi:10.1371/JOURNAL.PMED.1002085

14. Kim J, Kim M, Shin Y, Cho JH, Lee D, Kim Y. Association between dietary diversity score and metabolic syndrome in Korean adults: a community-based prospective cohort study. Nutrients. 2022;14. doi:10.3390/NU14245298/S1

15. Takeuchi K, Naito M, Kawai S, et al. Study profile of the Japan Multi-institutional Collaborative Cohort (J-MICC) Study. J Epidemiol. 2021;31:660–668. doi:10.2188/JEA.JE20200147

16. Tokudome S, Ikeda M, Tokudome Y, Imaeda N, Kitagawa I, Fujiwara N. Development of data-based semi-quantitative food frequency questionnaire for dietary studies in middle-aged Japanese. Jpn J Clin Oncol. 1998;28:679–687. doi:10.1093/jjco/28.11.679

17. Tokudome Y, Goto C, Imaeda N, et al. Relative validity of a short food frequency questionnaire for assessing nutrient intake versus three-day weighed diet records in middle-aged Japanese. J Epidemiol. 2005;15:135. doi:10.2188/JEA.15.135

18. Kant AK, Schatzkin A, Harris TB, Ziegler RG, Block G. Dietary diversity and subsequent mortality in the First National Health and Nutrition Examination Survey Epidemiologic Follow-up Study. Am J Clin Nutr. 1993;57:434–440. doi:10.1093/AJCN/57.3.434

19. de Oliveira Otto MC, Anderson CAM, Dearborn JL, et al. Dietary diversity: implications for obesity prevention in adult populations: a science advisory from the American Heart Association. Circulation. 2018;138:e160. doi:10.1161/CIR.0000000000000595

20. Azadbakht L, Mirmiran P, Esmaillzadeh A, Azizi F. Dietary diversity score and cardiovascular risk factors in Tehranian adults. Public Health Nutr. 2006;9:728–736. doi:10.1079/PHN2005887

21. Grundy SM, Cleeman JI, Daniels SR, et al. Diagnosis and management of the metabolic syndrome. Circulation. 2005;112:2735–2752. doi:10.1161/CIRCULATIONAHA.105.169404

22. Van Nguyen T, Arisawa K, Katsuura-Kamano S, et al. Associations of metabolic syndrome and metabolically unhealthy obesity with cancer mortality: The Japan Multi-Institutional Collaborative Cohort (J-MICC) study. PLoS One. 2022;17:e0269550. doi:10.1371/JOURNAL.PONE.0269550

23. Li P, Stuart EA, Allison DB. Multiple imputation: a flexible tool for handling missing data. JAMA. 2015;314:1966. doi:10.1001/JAMA.2015.15281

24. Oldewage-Theron WH, Kruger R. Food variety and dietary diversity as indicators of the dietary adequacy and health status of an elderly population in Sharpeville, South Africa. J Nutr Elder. 2008;27:101–133. doi:10.1080/01639360802060140

25. Tian X, Wu M, Zang J, Zhu Y, Wang H. Dietary diversity and adiposity in Chinese men and women: an analysis of four waves of cross-sectional survey data. European Journal of Clinical Nutrition 2017 71:4. 2016;71(4):506–511. doi:10.1038/ejcn.2016.212

26. Xu X, Inglis SC, Parker D. Sex differences in dietary consumption and its association with frailty among middle-aged and older Australians: a 10-year longitudinal survey. BMC Geriatr. 2021;21(1):1–12. doi:10.1186/S12877-021-02165-2/TABLES/4

27. Farhangi MA, Jahangiry L. Dietary diversity score is associated with cardiovascular risk factors and serum adiponectin concentrations in patients with metabolic syndrome. BMC Cardiovasc Disord. 2018;18:1–6. doi:10.1186/S12872-018-0807-3/FIGURES/1

28. Ogilvie RP, Lutsey PL, Heiss G, Folsom AR, Steffen LM. Dietary intake and peripheral arterial disease incidence in middle-aged adults: the Atherosclerosis Risk in Communities (ARIC) Study. Am J Clin Nutr. 2017;105:651. doi:10.3945/AJCN.116.137497

29. Luna-Castillo KP, Olivares-Ochoa XC, Hernández-Ruiz RG, et al. The effect of dietary interventions on hypertriglyceridemia: from public health to molecular nutrition evidence. Nutrients. 2022;14. doi:10.3390/NU14051104

30. Otsuka R, Tange C, Nishita Y, et al. Dietary diversity and all-Cause and cause-specific mortality in Japanese community-dwelling older adults. Nutrients. 2020;12. doi:10.3390/NU12041052

31. Miyamoto K, Kawase F, Imai T, Sezaki A, Shimokata H. Dietary diversity and healthy life expectancy—an international comparative study. European Journal of Clinical Nutrition 2018 73:3. 2018;73:395–400. doi:10.1038/s41430-018-0270-3

32. Salehi-Abargouei A, Akbari F, Bellissimo N, Azadbakht L. Dietary diversity score and obesity: a systematic review and meta-analysis of observational studies. European Journal of Clinical Nutrition 2016 70:1. 2015;70:1–9. doi:10.1038/ejcn.2015.118

